# Preferential preservation of visuospatial memory over verbal memory in the Old Order Amish

**DOI:** 10.1101/2021.07.06.21259942

**Authors:** Michael B. Prough, Laura J. Caywood, Jason E. Clouse, Sharlene D. Herington, Susan H. Slifer, Daniel A. Dorfsman, Larry D. Adams, Reneé A. Laux, Yeunjoo E. Song, Audrey Lynn, M. Denise Fuzzell, Sarada L. Fuzzell, Sherri D. Hochstetler, Kristy Miskimen, Leighanne R. Main, Michael D. Osterman, Paula Ogrocki, Alan J. Lerner, Jairo Ramos, Jeffery M. Vance, Michael L. Cuccaro, Jonathan L. Haines, William K. Scott, Margaret A. Pericak-Vance

## Abstract

**Background:** While studying cognition in the Old Order Amish (OOA), we have observed strong performance on the constructional praxis delayed recall (CPDR) as compared to other cognitive tests, independent of overall cognitive status. This may indicate a preferential preservation of visuospatial memory in this population. Here, we investigate this by comparing the CPDR to the word list delayed recall (WLDR) within the OOA, as well as by comparing these results to a non-Amish cohort.

**Method:** 420 OOA individuals in Indiana/Ohio age 66-95 who had complete data for the CPDR and WLDR were included. From the non-Amish CERAD cohort, 401 individuals age 60-96 with the same tests were included. For both cohorts, education-adjusted Z-scores were calculated for the CPDR and WLDR. The difference between the CPDR Z-score and the WLDR Z-score was calculated as a measure of the preservation of visuospatial memory over verbal memory. T-tests were first used to compare the tests within both cohorts and then stratified by case/control status. Linear regression was then used to investigate the effects of age, sex, cognitive status, and cohort on the Z-scores and difference between Z-scores. Additional t-tests and regressions were then performed to further investigate the effect of sex and its interaction with cohort.

**Result:** We found a significantly better performance on CPDR over WLDR in every cognitive status group in the OOA, but not in all groups of the CERAD cohort. After controlling for age, sex, and cognitive status, this preferential preservation remains significantly higher in the Amish, with being in the Amish cohort increasing the difference between Z-scores by an average of 0.615 units when compared to being in the CERAD cohort. When adjusting for age, sex, cognitive status, and cohort, the interaction between cohort and sex is significant, with the Amish males exhibiting a greater difference between Z-scores compared to other groups, with a significant interaction value of 0.676.

**Discussion:** Overall, these findings suggest that the OOA preferentially preserve visuospatial memory over verbal memory, regardless of cognitive status. This effect is particularly strong in OOA males. In summary, this study gives additional evidence that the Amish exhibit unique patterns of memory loss and aging, with a preferential preservation of visuospatial memory over verbal memory. Additional studies are needed to further explain this phenomenon.

## INTRODUCTION

One of the hallmark symptoms of Alzheimer disease (AD) and related dementias is impairment of memory (Borelli et al., 2020; Jahn, 2013). While older adults generally show a decline in memory and other cognitive abilities over time, there are vast differences in the amount and progression of decline characteristic of normal aging from the decline seen in AD and related disorders (Abichou et al., 2020; El Haj et al., 2020). Our current understanding is that memory is a multi-dimensional process and varies by content (e.g., verbal, procedural, visuospatial, etc), and characteristics related to encoding, storage, and retrieval. Interestingly, the various dimensions of memory appear to be specific to brain regions. As the brain structure and function changes during aging, memory performance is impacted (Preston & Eichenbaum, 2013; Sekeres et al., 2018). Depending on the location and extent of brain changes, there are distinct forms of memory loss, such as dementia. For example, degeneration of medial temporal and parietal regions of the brain lead to memory impairment in AD, whereas the same memory capabilities are intact in other forms of dementia, even when there is significant atrophy of hippocampal regions (Irish et al., 2016).

Consistent with the above findings, there is evidence that visuospatial memory is mediated by the entorhinal-hippocampal circuit of the brain (Eichenbaum, 1999; Eichenbaum et al., 2012; Knierim et al., 2014; Norman & Eacott, 2005; Staresina et al., 2011). This is thought to be one of the first sites of preclinical AD pathology (Haque et al., 2019), and recent studies have used sensitive visuospatial memory tasks, such as VisMET (Visuospatial Memory Eye-Tracking Task), as screening tools for memory impairment and preclinical AD. Visuospatial memory has been assessed using a variety of tasks, such as reproducing line drawings, replicating designs with blocks, etc. (Corey-Bloom et al., 2016; Shin et al., 2006). While many of these tasks have a motor component, the visuospatial memory tasks require the ability to identify details and structure of forms in multiple dimensions of space (Dickerson & Atri, 2014). A widely used visuospatial memory task is the delayed recall portion of Constructional Praxis (CP) test (Rosen et al., 1984) for which participants copy shapes and reproduce them from memory after a short delay. For the recall component, participants are asked to draw the four shapes from memory after a delay, during which other test are administered, and the shapes are again scored, with a total maximum score of 11 points.

In contrast to visuospatial memory, verbal memory involves different prefrontal and parietal areas of the brain (Binder et al., 2005; Cabeza et al., 1997; Grady et al., 1995; Klostermann et al., 2008). Typical tests for verbal memory include story recall and word list learning (Bowden et al., 2011; Wiens et al., n.d.). The CERAD battery contains the Word List (WL) test, which consists of immediate and delayed recall (Hankee et al., 2016). Comparison of males and females on visuospatial and verbal memory have found that women outperform men on verbal memory tasks, whereas men outperform women on memory tasks that require visuospatial processing (Herlitz, 2001; Herlitz et al., 1997; Herlitz & Rehnman, 2008). While sex differences between visuospatial and verbal memory are well studied, there is limited evidence of a preferential preservation of one type of memory over another. As part of an ongoing study of the Old Order Amish (OOA), we have cognitive data that allows us to examine preferential preservation in this group.

The OOA are a relatively genetically homogeneous population that came to the United States in the eighteenth and nineteenth centuries (Hahs et al., 2006; van der Walt et al., 2005). There are now large settlements of OOA in Pennsylvania, Indiana, and Ohio, among other areas of the country. This population not only exhibits genetic homogeneity but also homogeneity in lifestyle traits, as they adhere to a similar set of religious beliefs and practices, such as abstaining from some modern conveniences such as electricity and automobiles (Bassett et al., 2004). Additionally, many OOA hold similar primary occupations, such as farming or housewifery. Given this relative homogeneity, multiple studies have investigated the genetics of cognitive disorders of aging in the OOA (Ashley-Koch et al., 2005; Cummings et al., 2012; D’Aoust et al., 2015; Hahs et al., 2006; Holder & Warren, 1998; Pericak-Vance et al., 1996). During these studies, we observed better performance on the Constructional Praxis Delay test as compared to other cognitive tests, especially in men, independent of overall cognitive status. This preferential preservation of visuospatial memory has not been well studied in the Amish or other populations.

Based on these observations, we compared visuospatial memory performance (using the Constructional Praxis Delay) to verbal memory performance (using Word List Delay) in both Amish and non-Amish populations of cases and controls. Our primary hypotheses are that (a) the OOA will exhibit a preferential preservation of visuospatial memory relative to verbal memory, and (b) this pattern will be unique to the OOA population.

## METHODS

### Study Populations

#### Collaborative Amish Aging and Memory Project (CAAMP)

Participants were drawn from ongoing studies of aging and memory in the OOA communities located in Adams, Elkhart, and LaGrange counties in Indiana and Holmes County in Ohio. Study participants were ascertained as previously described (Pericak-Vance et al., Hahs et al., and Edwards et al.). All participants were administered a standard battery of neuropsychological tests including the Constructional Praxis Delay and the Word List Delay tests (Edwards et al., 2011, 2013; Hahs et al., 2006; Pericak-Vance et al., 1996), which were used for categorization of cognitive status by an adjudication panel. Blood samples for DNA and RNA extraction were also collected, and genome-wide genotype data were generated from these samples using a Multi-Ethnic Global Array (MEGA) or Global Screening Consortium (GSA) array from Illumina.

#### CERAD Normative Cohort

The Consortium to Establish a Registry for Alzheimer’s Disease (CERAD) enrolled participants with and without Alzheimer disease from 1987 to 1995. Participants underwent clinical evaluations that included a neuropsychological assessment; the battery consisted of multiple tests including the Constructional Praxis Delay and Word List Delay tests (Fillenbaum et al., 2008). In this study, the CERAD cohort serves as a non-Amish comparison to the Amish cohort, as the two cohorts share common variables such as age, sex, education, cognitive status (case vs control), and both the Constructional Praxis Delay and Word List Delay tests (Morris et al., 1989).

### Measures

#### Constructional Praxis-Delay (CP) (Morris et al., 1989)

The CP test consists of a copy portion in which participants copy four shapes (circle, diamond, overlapping rectangles, and a cube). Reproductions are scored based on accuracy using well-defined criteria, with a total maximum score of 11 points. For the CERAD battery, the copy portion was expanded to include a delayed recall component to better characterize visuospatial memory (Fillenbaum et al., 2008, 2011). For the recall component, participants are asked to draw the four shapes from memory after a delay, and the shapes are again scored, with a total maximum score of 11 points.

#### CERAD Word List Learning test (WLL)

(Hankee et al., 2016; Morris et al., 1989). The WLL is a 10-item word list which is read aloud to participants who are then asked to immediately recall the words over three trials and, following a delay to recall as many of these ten words as they can. The delay trial score is the number of words correctly recalled.

### Data Processing

From the Amish cohort, we analyzed data from 420 individuals age 66-95 who had complete data for the Constructional Praxis Delay and the Word List Delay tests on the same date, had genome-wide genotype data, and had a case conference categorization of Cognitively Impaired (cases) or Cognitively Unimpaired (controls). Participants categorized as Borderline were excluded from analysis. From the CERAD cohort, we analyzed data from 401 individuals age 60-96 who had complete data for the Constructional Praxis Delay and the Word List Delay tests on the same date and were classified as AD cases or cognitively unimpaired controls.

For both cohorts, education-adjusted Z-scores were calculated for the Constructional Praxis Delay test, using 10-year age group normative data from the Cache County Memory study (Fillenbaum et al., 2011; Welsh-Bohmer et al., 2009). Education-adjusted Z-scores were also calculated for the Word List Delay test, using 20-year age group normative data derived from 23 US CERAD sites and participants enrolled in an Alzheimer’s Disease Research Center study (Beeri et al., 2006; Welsh et al., 1994). The difference between the Constructional Praxis Delay Z-Score and the Word List Delay Z-Score was calculated as a measure of the preservation of Constructional Praxis Delay over the Word List Delay.

### Statistical Analysis/Data Analysis

Our preliminary analyses compared individual performances on the Constructional Praxis Delay Z-score and the Word List Delay Z-score using paired t-tests in both the Amish and CERAD cohorts, first including all participants, and then stratifying into separate case and control categories. This was followed by comparison of mean Constructional Praxis Delay Z-score, Word List Delay Z-score, and difference between these Z-scores between our Amish and CERAD cohorts using independent two-sample t-tests. Similarly, these independent two-sample t-tests first included all participants, then stratified into separate case and control categories. We then investigated the effects of age, sex, cognitive status, and cohort (Amish vs CERAD) on the Constructional Praxis Delay Z-score, Word List Delay Z-score, and difference between these Z-scores using multiple linear regression. A kinship matrix was calculated in the Amish cohort using genome-wide genotype data in order to adjust for relatedness within the model. Because the effect of these covariates on the outcome variables seems to differ by cognitive status, we also stratified this linear regression into case and control categories.

Additionally, to further investigate the effect of sex on the outcomes of interest we performed additional t-tests in subgroups by sex and cohort in order to highlight these differences (Amish males vs Amish females and CERAD males vs CERAD females). Finally, we performed additional multiple linear regressions that included an interaction term between sex and cohort. All t-tests were conducted using the Statistical Analysis System 9.3 (SAS 9.3) and regressions and kinship matrix calculations were conducted using the GENESIS program in RStudio 1.3.959.

## RESULTS

Demographic information for each cohort is presented in Table 1. Compared to the CERAD cohort, the Amish cohort contains a higher proportion of controls to cases (66.4% vs 42.9%) and has an older average age (81.9 vs 75.2). The proportion of females to males is similar between the two cohorts (61.7% vs 61.3%).

**Table 1.** Demographic information for each cohort

| Table 1. Demographic information for each cohort |  |  |
| --- | --- | --- |
|  | Amish | CERAD |
| Total N | 420 | 401 |
| Cases | 141<br>(33.6%) | 229<br>(57.1%) |
| Controls | 279<br>(66.4%) | 172<br>(42.9%) |
| Average Age | 81.9 | 75.2 |
| Age Range | 66-95 | 60-96 |
| Percent Female | 61.7% | 61.3% |

T-test results show a significant preferential preservation of Constructional Praxis Delay over Word List Delay in all Amish groups at p<0.0001 (Figure 1). In the CERAD cohort, this preferential preservation is only observed in the cases group, but with a smaller effect size than in the corresponding Amish cases group (difference in mean Z-score of 1.61 in the Amish cases vs 0.36 in the CERAD cases). Notably, in the CERAD controls group, the Word List Delay Z-score is significantly higher than the Constructional Praxis Delay Z-score, showing the reverse direction of the preferential preservation seen in the Amish groups, with the Word List Delay being slightly preserved over the Constructional Praxis Delay in this CERAD controls group.

**Figure 1.**
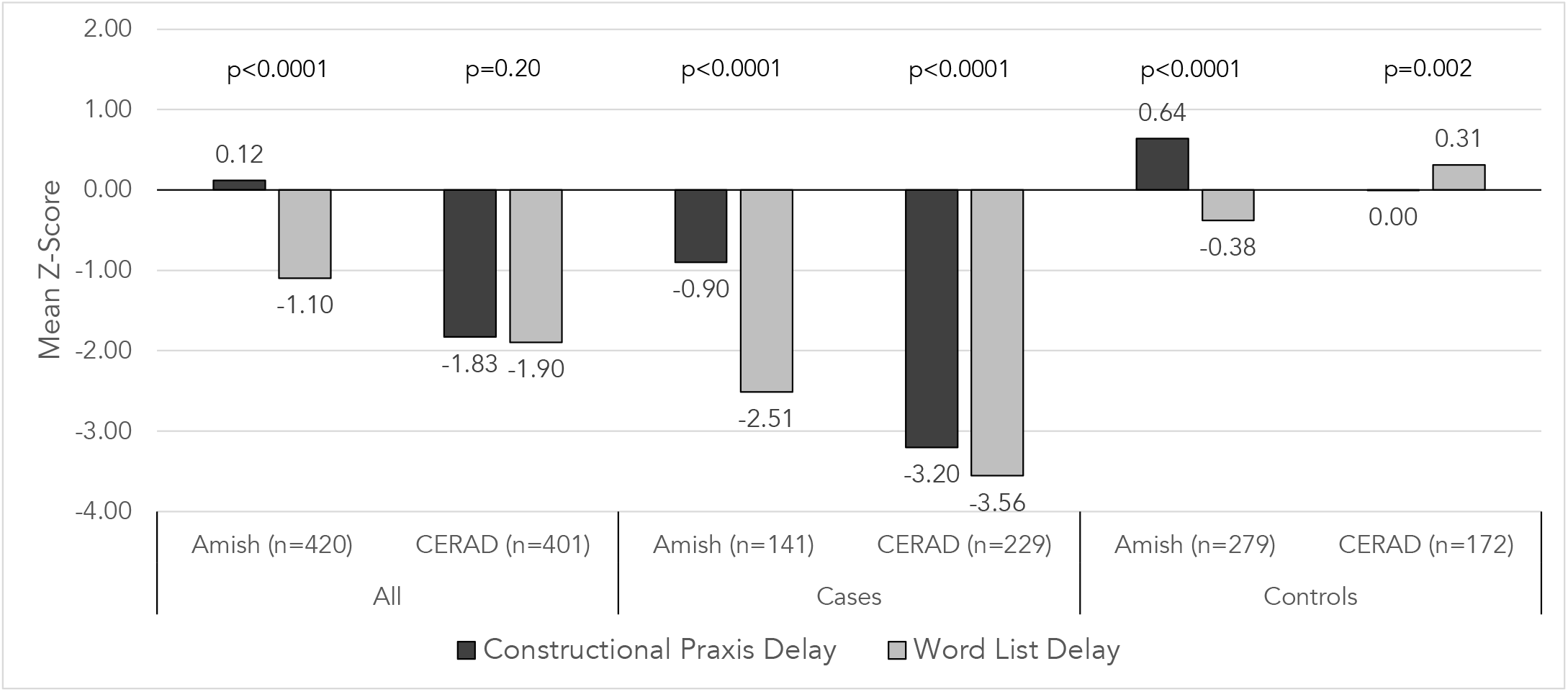
Mean Constructional Praxis Delay Z-score compared to mean Word List Delay Z-score by cohort and cognitive status.

T-tests comparing this preferential preservation between the Amish and CERAD cohorts show significantly higher preservation in the Amish, both when the entire sample is considered (1.22 vs 0.07), and also when stratified into cases (1.61 vs 0.35) and controls (1.02 vs -0.31), all at p-values of <0.0001 (Figure 2). After controlling for age, sex, and cognitive status, this preferential preservation remains significantly higher in the Amish, with being in the Amish cohort increasing the difference between Z-scores by an average of 0.615 units when compared to being in the CERAD cohort, at a p-value of <0.0001 (Table 2). This is further illustrated when looking at the Constructional Praxis Delay and Word List Delay test separately, where being Amish significantly increases the Constructional Praxis Delay Z-score but does not significantly increase the Word List Delay Z-Score (Table 2). This suggests that the preferential preservation of the Constructional Praxis Delay in the Amish is a result of a relative preservation of Constructional Praxis Delay as opposed to poorer performance on Word List Delay. Furthermore, when these results are stratified into case and control groups, when adjusting for age and sex, the preferential preservation increases going from the CERAD to the Amish cohort in both the cases and control groups (Table 3). Being Amish increases the difference between Z-scores by 1.097 in the cases and 1.154 in the controls.

**Table 2.**
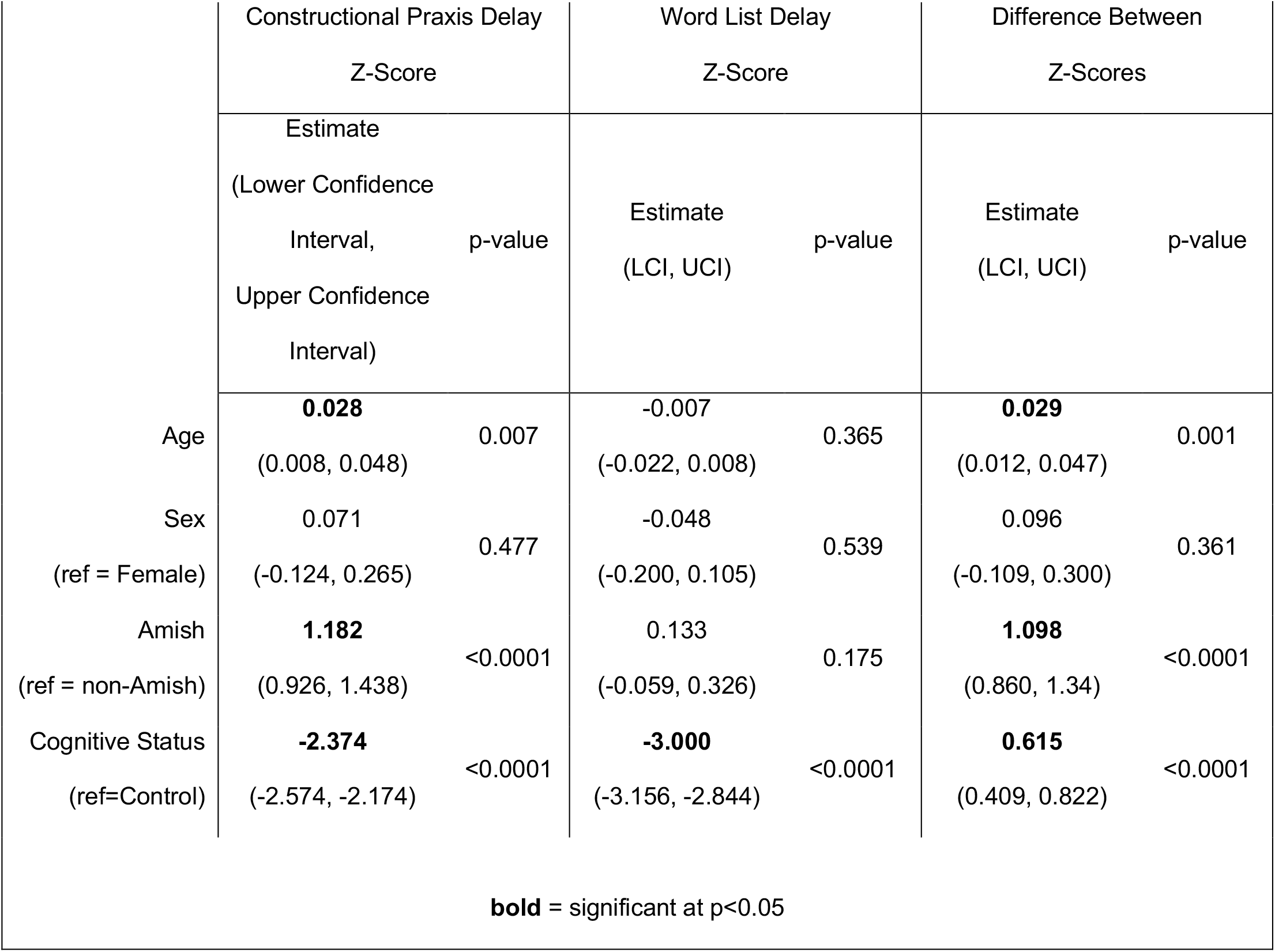
Adjusted multivariate linear regression of Constructional Praxis Delay Z-score, Word List Delay Z-score, and difference between these Z-scores

**Table 3.**
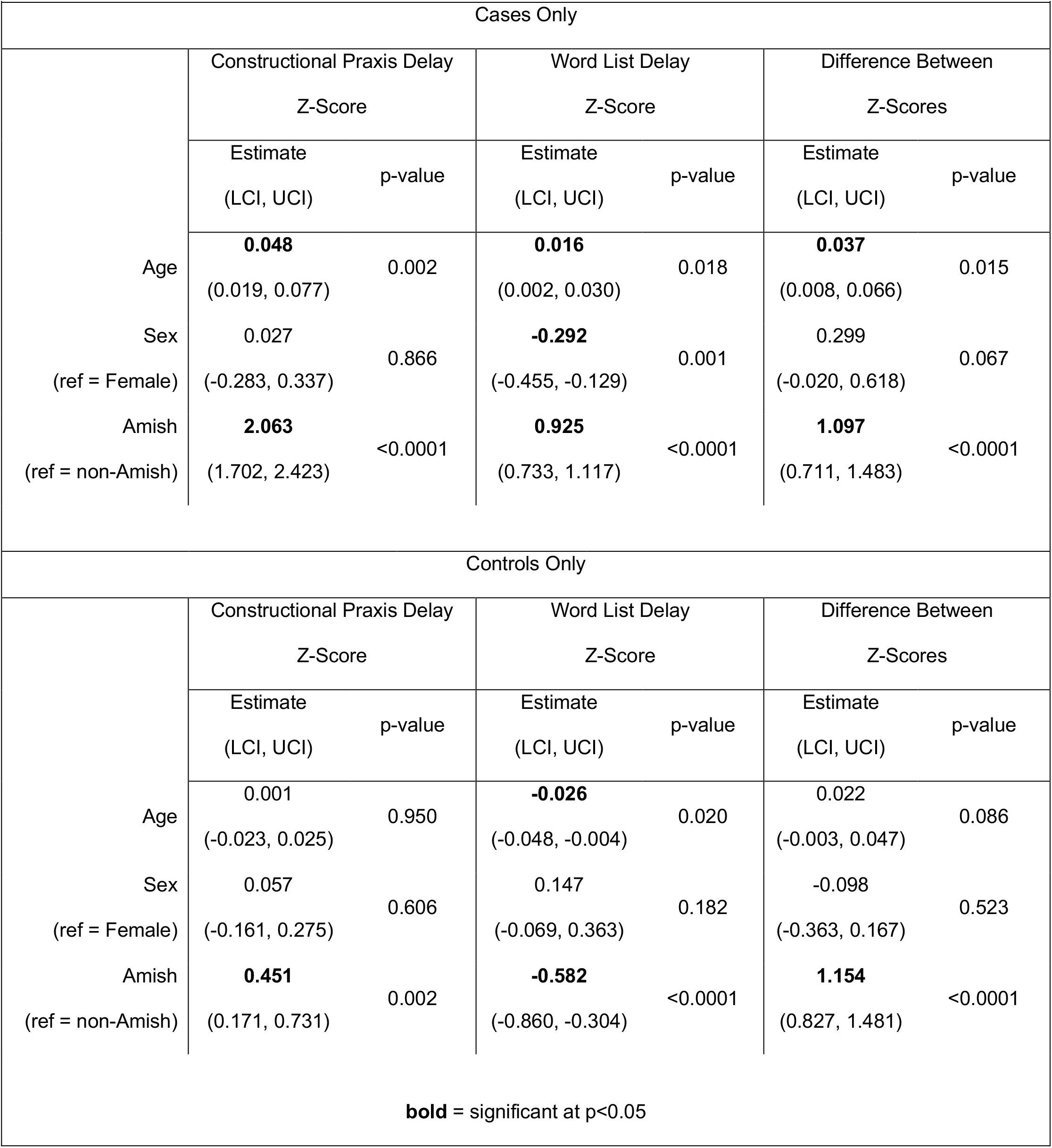
Linear regression of Constructional Praxis Delay Z-score, Word List Delay Z-score, and difference between these Z-scores stratified by cognitive status

**Figure 2.**
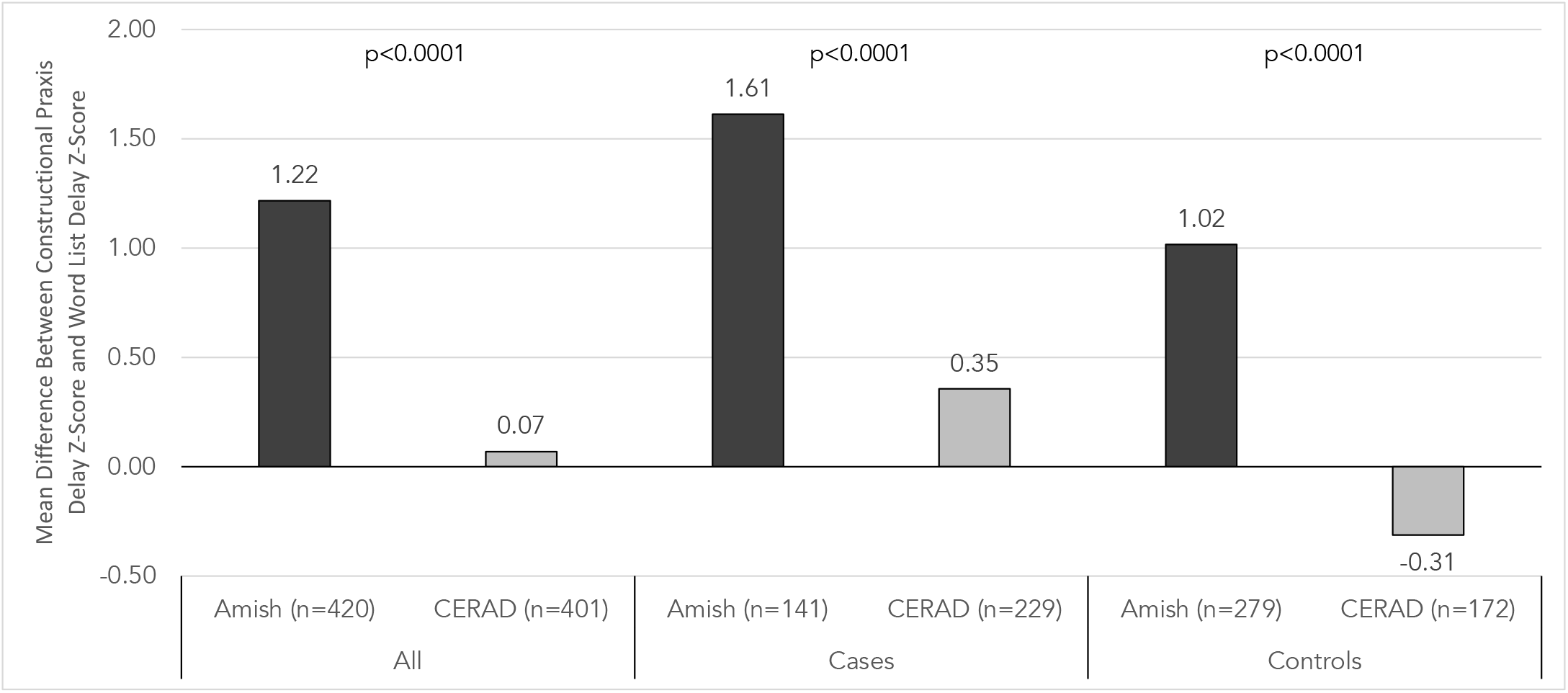
Mean difference between Constructional Praxis Delay Z-score and Word List Delay Z-score by cohort and cognitive status.

T-test results comparing differences in Constructional Praxis Z-score to Word List Delay Z-score between males and females show that in the Amish cohort, this preferential preservation is significantly higher in the males. In the CERAD cohort, a slight preferential preservation is only seen in the females but not in the males, showing that this effect is opposite in the CERAD cohort (Figure 3). Additionally, when adjusting for age, sex, cognitive status, and cohort, the interaction between cohort and sex is significant, with the Amish males exhibiting a greater difference between Z-scores compared to other groups, with a significant interaction value of 0.676 (Table 4). Furthermore, when looking at the Constructional Praxis Delay and Word List Delay tests separately in this model, this interaction term is significant in the Constructional Praxis Delay but not the Word List Delay, suggesting that the increased preferential preservation of the Constructional Praxis Delay in Amish males is a result of a relative preservation of Constructional Praxis Delay as opposed to a reduction in Word List Delay when compared to other groups.

**Table 4.**
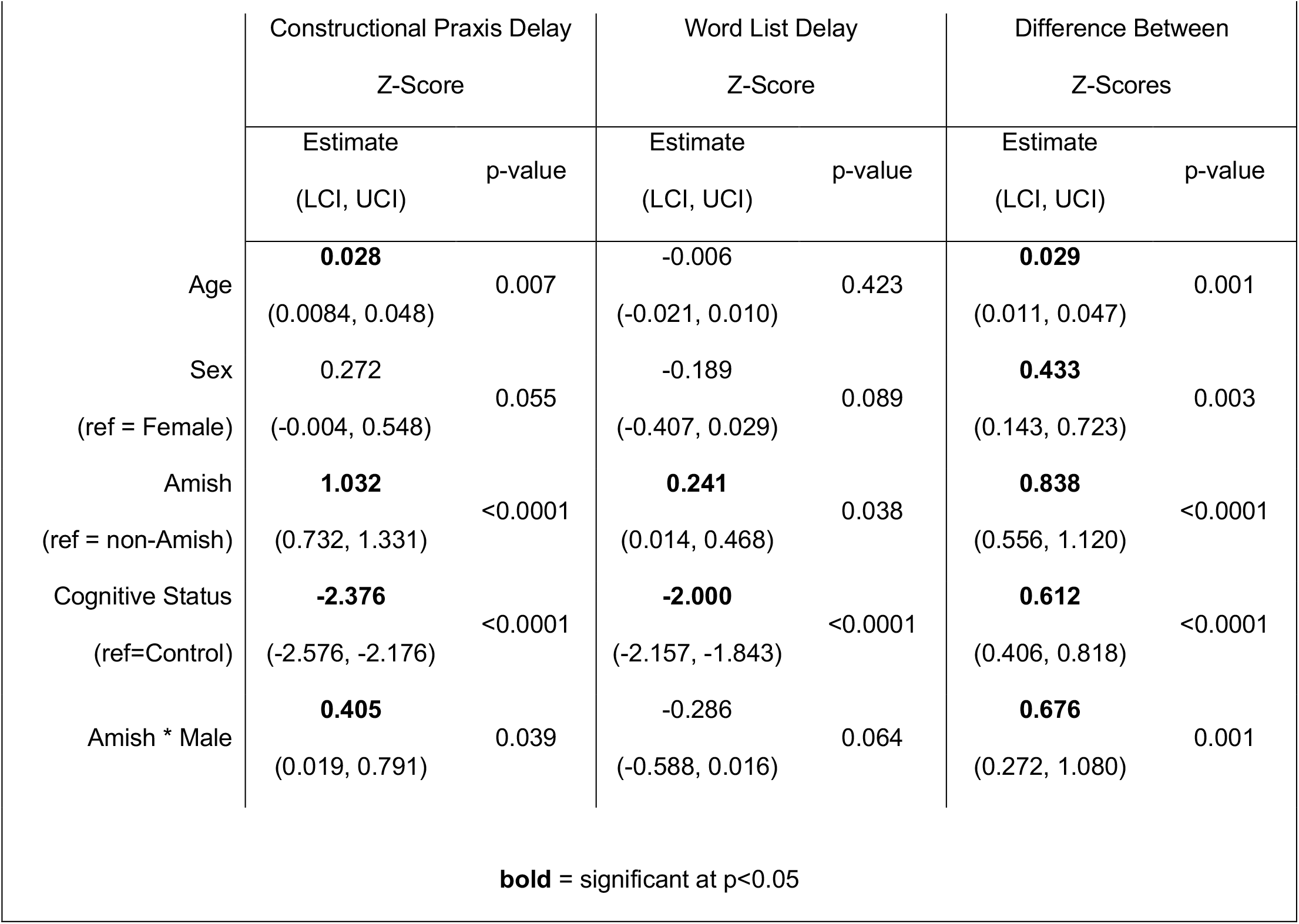
Linear regression of Constructional Praxis Delay Z-score, Word List Delay Z-score, and difference between these Z-scores with Amish * Male interaction

|  | Constructional Praxis Delay<br>Z-Score |  | Word List Delay<br>Z-Score |  | Difference Between<br>Z-Scores |  |
| --- | --- | --- | --- | --- | --- | --- |
|  | Estimate<br>(LCI, UCI) | p-value | Estimate<br>(LCI, UCI) | p-value | Estimate<br>(LCI, UCI) | p-value |
| Age | **0.028**<br>(0.0084, 0.048) | 0.007 | -0.006<br>(-0.021, 0.010) | 0.423 | **0.029**<br>(0.011, 0.047) | 0.001 |
| Sex<br>(ref = Female) | 0.272<br>(-0.004, 0.548) | 0.055 | -0.189<br>(-0.407, 0.029) | 0.089 | **0.433**<br>(0.143, 0.723) | 0.003 |
| Amish<br>(ref = non-Amish) | **1.032**<br>(0.732, 1.331) | <0.0001 | **0.241**<br>(0.014, 0.468) | 0.038 | **0.838**<br>(0.556, 1.120) | <0.0001 |
| Cognitive Status<br>(ref=Control) | **-2.376**<br>(-2.576, -2.176) | <0.0001 | **-2.000**<br>(-2.157, -1.843) | <0.0001 | **0.612**<br>(0.406, 0.818) | <0.0001 |
| Amish * Male | **0.405**<br>(0.019, 0.791) | 0.039 | -0.286<br>(-0.588, 0.016) | 0.064 | **0.676**<br>(0.272, 1.080) | 0.001 |
| **bold** = significant at p<0.05 |  |  |  |  |  |  |

**Figure 3.**
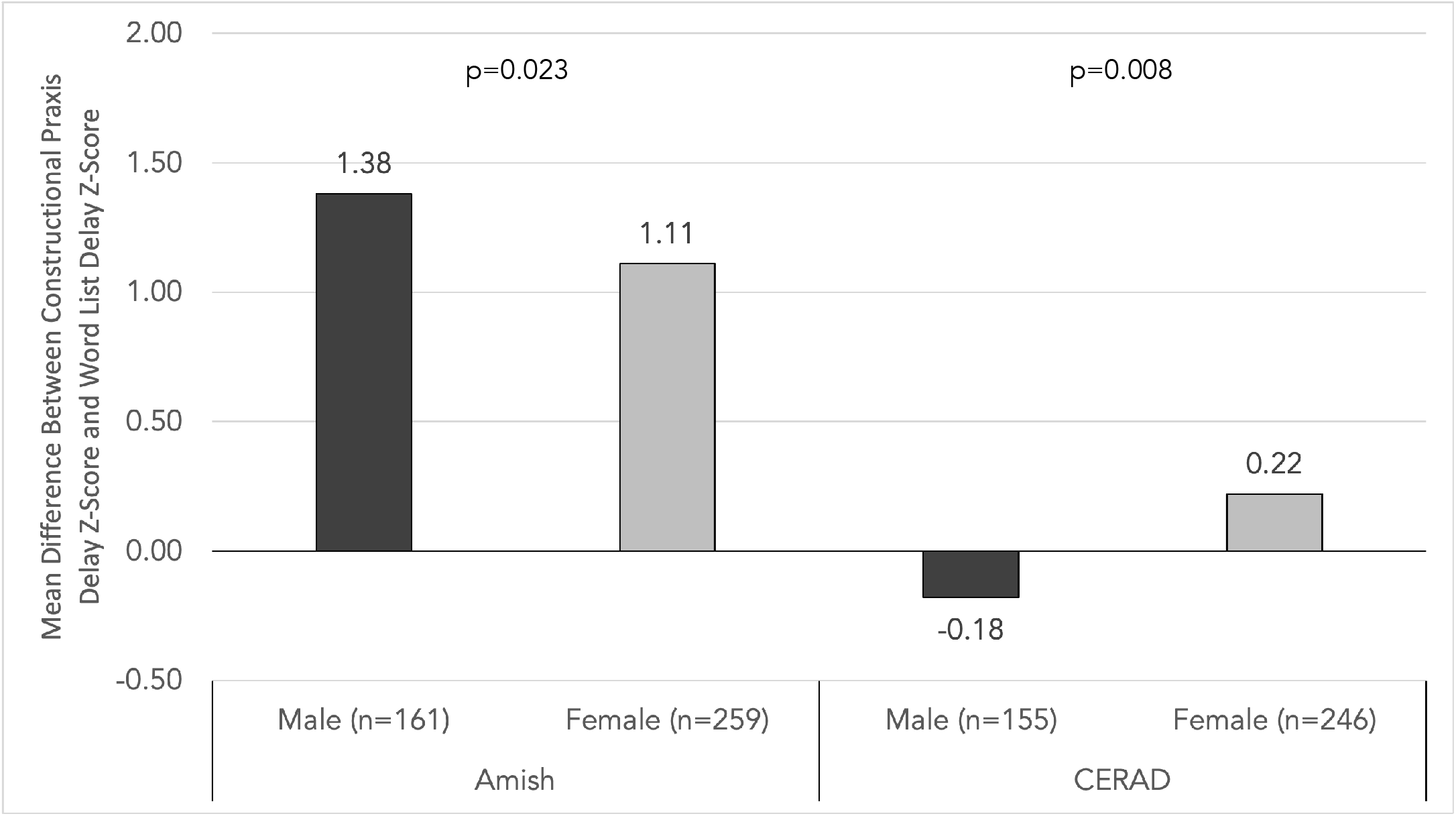
Mean difference between Constructional Praxis Delay Z-score and Word List Delay Z-score by cohort and sex.

## DISCUSSION

Based on the multivariate linear regression models, our results show that the OOA exhibit a greater difference between Constructional Praxis Delay Z-score and Word List Delay Z-score than the CERAD cohort, when adjusting for age, sex, and cognitive status. Furthermore, when stratified by cognitive status, we see that this difference is significant and of a similar magnitude in both the cases and controls.

When further investigating the interaction between sex and cohort in this study, our results show that being male and Amish has an interactive effect, suggesting that Amish males have the greatest difference between Constructional Praxis Delay and Word List Delay tests when compared to other groups in the study and that this difference is due to a preservation of Constructional Praxis Delay rather than a decrease in Word List Delay. This is consistent with previous studies that have found that women outperform men on verbal memory tasks, whereas men outperform women on memory tasks that require visuospatial processing (Herlitz, 2001; Herlitz et al., 1997; Herlitz & Rehnman, 2008)

Overall, these findings suggest that the OOA preferentially preserve visuospatial memory over verbal memory, which could be the result of different influences. For example, while one could speculate about possible neurobiological differences, it may be that performance on visuospatial tasks, which engage specific cognitive skills, are bolstered by well-learned life experiences and often show little decline (Blazer et al., 2015). As such, the sparing of visuospatial abilities could be the result of reserve. Given the relative homogeneity of life and occupational experience we would expect visuospatial abilities to be spared. At the same time, impaired visuospatial abilities (e.g., poor performance on the CP test) could be a sensitive indicator of decline when testing individuals at multiple timepoints.

In the future, we will further investigate this phenomenon by comparing the Constructional Praxis Delay to other tests in the neuropsychological battery to see if this pattern holds when compared to other cognitive processes. In addition, we will investigate both genetic and non-genetic factors that may explain this pattern in the Amish, especially in men. Non-genetic factors of interest include more detailed occupational, lifestyle, and non-formal educational information, as we suspect specific occupations and hobbies that are common among Amish males (e.g., woodworking, farming, or others that are highly visuospatial) could play a role in this phenomenon. These occupations and life experiences could allow for additional non-formal education and practice with visuospatial tasks, resulting in a cognitive reserve specific to these visuospatial processes. This would be similar to evidence that links years of formal education with cognitive reserve, which protects against neurodegenerative disease (Farfel et al., 2013; Lövdén et al., 2020; Luerding et al., 2016; Ramos et al., 2021).

One limitation of this study is the relatively small sample sizes that included the necessary data in both the Amish and CERAD cohorts. Another limitation is the absence of imaging data such as MRI results in any of the Amish participants, which limits the ability to make a definitive diagnosis of AD in this population. If imaging results had been available, they may have been compared to imaging in the CERAD group to further investigate function in specific regions of the brain associated with these different types of memory.

In summary, this study gives additional evidence that the Amish exhibit unique patterns of memory loss and aging, with a preferential preservation of visuospatial memory over verbal memory. Additional studies are needed to further explain this phenomenon.

## Data Availability

Data referred to in the manuscript may be obtained with permission of the study PIs by contacting the primary author.

## ACKNOWLEDGEMENTS

We thank the Amish families for allowing us into their communities and for participating in our study. We also acknowledge the contributions of the Anabaptist Genealogy Database and Swiss Anabaptist Genealogy Association. This study is supported by National Institutes of Health / National Institute of Aging, grant 1RF1AG058066 (to Jonathan L. Haines, Margaret Pericak-Vance, and William K. Scott). Finally, we acknowledge the resources provided by the Department of Population and Quantitative Health Sciences, School of Medicine at Case Western Reserve University and the John P. Hussman Institute for Human Genomics at University of Miami, Miller School of Medicine.

